# Universal PCR for Bacteria, Mycobacteria, and Fungi: A 10-year Retrospective Review of Clinical Indications and Patient Outcomes

**DOI:** 10.1101/2023.08.02.23293145

**Authors:** Jeffrey Kubiak, Alexandra Morgan, Andrea Kirmaier, Ramy Arnaout, Stefan Riedel

## Abstract

Universal PCR for bacteria, mycobacteria, and fungi can aid in the diagnosis of occult infections, especially in the case of fastidious organisms or when prior antimicrobial treatment compromises culture growth. However, the limitations of this technology, including lack of specificity, high cost, long turnaround time, and lack of susceptibility data, may limit its effect on clinical outcomes. We performed a retrospective analysis of all specimens sent for universal PCR over a 10-year period from 2013-2022, focusing on clinical indications for test utilization and patient outcomes. A total of 708 specimens were sent from 638 patients. Of those specimens, 163 were positive, for an overall positivity rate of 23%. Pre-analytic factors associated with a positive universal PCR result were the presence of organisms on Gram stain or histology, the presence of neutrophils on Gram stain, and growth on culture. Positivity rates varied significantly by specimen type. 20% of all organisms detected were deemed contaminants by the clinical services. A positive universal PCR led to a change in antibiotic management in 29% of cases. Positive fungal universal PCR results sent from hospitalized patients were associated with worse outcomes, including increased hospital mortality. Our findings suggest that factors such as the presence of organisms or neutrophils on Gram stain, specimen source/clinical context, and anticipated changes in management based on results should be strongly considered when making stewardship decisions regarding the appropriateness of this testing modality.

## Introduction

Universal PCR for bacteria, mycobacteria, and fungi is an attractive technology for the diagnosis of occult infections that may not be detectable by routine microbiology laboratory methods^1^. As molecular diagnostics such as these continue to evolve, laboratory stewardship is becoming increasingly important to ensure that these novel technologies are being utilized to maximize benefit to the patient while minimizing unnecessary healthcare costs.

Universal PCR is best utilized in cases of culture-negative infections, especially in cases of fastidious organisms or when antimicrobials were given prior to specimen collection that compromise culture growth^2^. While universal PCR has a fairly low sensitivity as a standalone assay^3^, prospective studies have shown that when used in conjunction with culture, the overall detection rate may be enhanced over the use of culture alone^4–7^. Various studies have supported the use of universal PCR in cases of meningitis^5, 8, 9^, endophthalmitis^10, 11^, joint and bone infections^6, 12–15^, pleural effusions^16, 17^, fungal sinusitis,^18^ and endocarditis^19–22^. While positive universal PCR results have been reported to lead to changes in antimicrobial treatment, there is considerable variability between studies regarding the extent to which these results impact antimicrobial management^23–25^.

Universal PCR is not without its limitations. Due to its non-specific nature, there is a high rate of false-positive results from contamination, especially in non-sterile body sites^26^. Universal PCR also cannot provide antimicrobial susceptibility data, and even in the case of a positive result clinicians must rely on epidemiologic data to guide coverage. In the absence of in-house universal PCR testing capacity, sending specimens to reference laboratories can take a week or longer to return results to the ordering institution, leaving clinicians to rely on empiric coverage of suspected organisms during a critical time in hospitalized patients’ care. Lastly, this assay comes at a high cost compared to other routine microbiology testing methods, typically hundreds of dollars^27^. Some studies have suggested that the combination of the lack of susceptibility data, the high rate of contamination, the sensitivity and faster turnaround time of concurrently submitted routine microbiology assays, and the cost mean that universal PCR provides little to no value to patient management or prognosis^28–30^.

In this context, we aimed to further address the clinical utility of universal PCR, focusing on patterns of utilization and patient outcomes. We performed a retrospective review of all universal PCR tests sent from our hospital network over a 10-year period and determined the impact of positive universal PCR results on patient antimicrobial management and hospitalized patient outcomes. These data may impart diagnostic and prognostic significance for clinicians to determine the merit of testing for their patients and can ultimately guide stewardship decisions about the use of this technology for challenging diagnostic cases.

## Methods

We performed a retrospective chart review for all patients for whom universal PCR testing was ordered from our 743-bed tertiary-care medical center. These orders were placed by both outpatient and inpatient clinical teams, typically in conjunction with the infectious disease consult service. Clinicians had the option to order any combination of bacterial, mycobacterial, or fungal universal PCR based on their clinical judgement of potential etiologies of the patient’s infection. All universal PCR orders required review and approval by a clinical director of our institution’s microbiology laboratories. The specimens were sent to the University of Washington Molecular Diagnosis Microbiology Section (Seattle, WA). Their test relies on sequencing of the 16S rRNA gene, the 65-kD heat shock protein, or the 28S rRNA gene plus internal transcribed spacer regions to broadly detect and speciate bacterial, mycobacterial, and fungal DNA sequences, respectively. In the case of bacterial and fungal universal PCR, reflex testing to next-generation sequencing was included in cases where multiple templates were present.

We queried our institution’s clinical data repository to retrieve information on all specimens ordered for universal PCR from our electronic medical record network over the 10-year period from January 2013 through November 2022. A chart review was performed on all patients to determine patient factors and outcomes, including demographic data, relevant clinical laboratory data including concurrent Gram stain results and culture results, specimen type, clinical indications, antibiotic treatment decisions, inpatient status, and hospitalization outcomes. We considered other laboratory tests to be concurrent with the specimen sent for universal PCR if either the same specimen was utilized for both assays or if both specimens were retrieved from the same location during the same surgical procedure. We relied on clinical judgement by physicians treating the patient to determine if a detected organism was considered clinically relevant or a presumed contaminant.

All analyses were performed using R 3.5.1 (R Foundation for Statistical Computing, Vienna, Austria) and RStudio 1.1.463 (Posit, Boston, MA). Population proportion confidence intervals were calculated to determine 95% confidence intervals for PCR positivity rates for different populations. A chi-squared test, odds ratio, or unpaired Student’s T-test was used for comparisons. P values were then corrected for multiple comparisons using the Benjamini-Hochberg method and a corrected p value of <0.05 was considered to be statistically significant. The Beth Israel Deaconess Medical Center Institutional Review Board reviewed the study protocol and determined it to be exempt (protocol no. 2017D000478).

## Results

Over the 10-year period from January 2013 to November 2022, a total of 708 specimens from 638 patients were sent to our reference laboratory for universal PCR. Of those, 574 were tested for bacteria, 321 for mycobacteria, and 294 for fungi. Universal PCR specimens had an average turnaround time of 12 days from order to result. The overall positivity rate was 23% (163/708, 95% CI 20-26%). The positivity rate for those tested for bacteria was 19% (109/574, 95% CI 16-22%), for mycobacteria was 6% (18/321, 95% CI 3.1-8.1%), and for fungi was 14% (40/294, 95% CI 9.7-18%) (**Figure 1A**).

**Figure 1.**
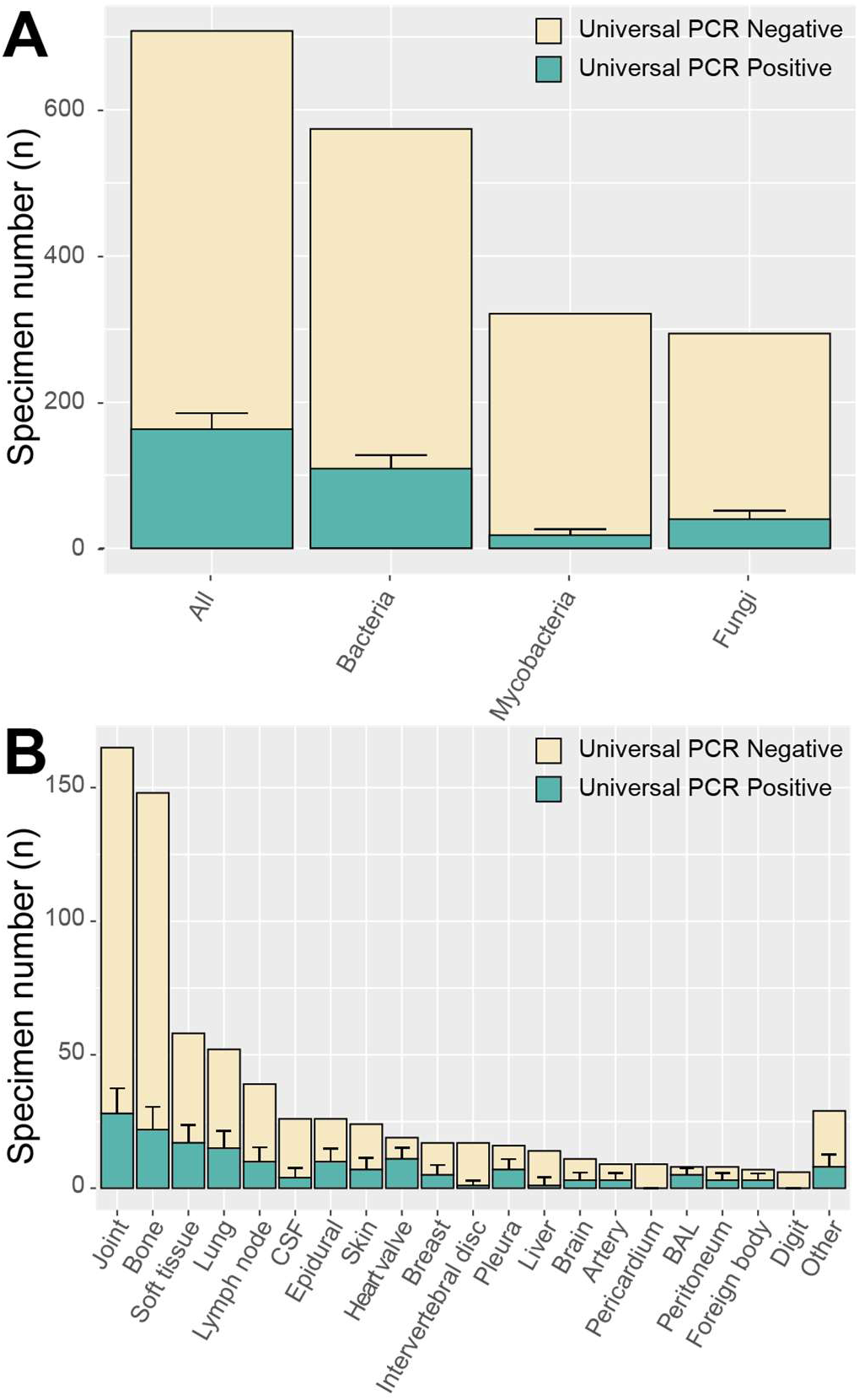
Total number of specimens sent for universal PCR and positivity rate by pathogen type (A) and by specimen source (B). A single specimen could be sent for more than one universal PCR at a time. Error bars represent the 95% population proportion confidence intervals for positive results.

We examined whether other laboratory tests performed on specimens sent for universal PCR were associated with a positive PCR result (**Table 1**), including Gram stain results and results of concurrent histopathology slides examined in surgical pathology. The presence of organisms on Gram stain (23/44 positive, 52% [95% CI 38-67%]) was associated with positive universal PCR results (p<0.05) and the absence of neutrophils on Gram stain (29/207 positive, 14% [95% CI 9-19%]) was negatively associated with positive universal PCR results (p<0.05). We observed no significant difference in positivity rate between paraffin-embedded blocks and fresh frozen tissue. However, the presence of organisms seen on histology slides (34/73 positive, 47% [95% CI 35-58%]) was also significantly associated with a positive universal PCR result (p<0.05). The presence of granulomata on histology was not associated with a positive PCR result.

**Table 1.** Pre-analytic determinants of positive universal PCR results.

|  | **Total**<br>**specimens** | **Total positives by**<br>**universal PCR** | **Percent positive**<br>**(95% CIs)** | **Corrected**<br>**p value** |
| --- | --- | --- | --- | --- |
| **Total** | 708 | 163 | 23% (20-26%) | --- |
| **Female Gender** | 342 | 73 | 21% (17-26%) | 0.81 |
| **Age > 50** | 498 | 113 | 23% (19-26%) | 0.97 |
| **Hardware Present** | 160 | 35 | 22% (15-28%) | 0.97 |
| **Gram stain organisms present** | **44** | **23** | **52% (38-67%)** | **0.0001** |
| **Neutrophils absent on gram**<br>**stain** | **178** | **29** | **16% (11-22%)** | **0.021** |
| **Cultures positive** | **154** | **53** | **34% (27-42%)** | **0.016** |
| **Organisms seen on histology** | **73** | **34** | **47% (35-58%)** | **0.0001** |
| **Granuloma on histology** | 72 | 12 | 17% (8-25%) | 0.42 |
| **Formalin fixed tissue** | 68 | 22 | 32% (21-43%) | 0.20 |
| **Outpatient** | 115 | 27 | 23% (16-31%) | 0.97 |
| **Inpatient** | 593 | 136 | 23% (20-26%) | 0.97 |

Specimens sent for universal PCR had concurrent cultures taken in 656 cases (93%), with 154/656 (23%) of those cultures positive for an organism. Having a culture positive for any organism was significantly associated with positive universal PCR results (53/154 positive, 34% [95% CI 27-42%], p<0.05). Of those cases with a positive culture and positive universal PCR result, there was concordance between those results for organism identification in 36/53 (68%) of cases.

A variety of specimen types were sent for universal PCR analysis. These included: artery, bronchoalveolar lavage, bone, brain (including dura and meninges), breast, cerebrospinal fluid, digits, intervertebral disc, epidural tissue, foreign bodies, joints, liver, lung, lymph node, pericardium, peritoneum, pleura, skin, soft tissue/muscle, heart valves, vitreous, esophagus, fallopian tube, bowel, bone marrow, uterus, and myocardium. Joint and bone tissue specimens were most frequent, with 165 and 148 specimens sent, respectively. Specimen types varied considerably in the overall positivity rates (**Figure 1B, Supplemental Table 1**). The highest positivity rates were seen in bronchoalveolar lavage specimens (5/8, 63% [95% CI 29%-96%]) and heart valves (11/19, 58% [95% CI 36%-80%]), and the lowest positivity rate was seen with pericardial specimens (0/9, 0%).

Likewise, clinical indications for testing varied considerably. The clinical indications for testing included: osteomyelitis, septic arthritis, abscess, granulomata seen on surgical pathology, mass/lesion, endocarditis, pneumonia, cellulitis, lymphadenopathy, pericardial or pleural effusion, arteritis, meningitis, peritonitis, sinusitis, aplastic anemia, endophthalmitis, or a non-healing surgical wound. The highest positivity rate was noted for endocarditis (11/23 positive, 48%, [95% CI 27-68%]). A complete table of positivity rates for specimen sources broken down by clinical indication and pathogen type can be found in **Supplemental Table 1**.

76% of all patients with positive universal PCR results were on empiric antimicrobial therapy prior to receiving results of the assay. Following the reporting of universal PCR results to the online medical record, clinicians considered the universal PCR result to be a contaminating organism in 33/163 (20%, 95% CI 14-26%) cases (**Figure 2A**). When broken down by specimen type, bronchoalveolar lavage (BAL) specimens were considered to have the highest contamination rate, with 4/5 (80%, 95% CI 45-100%) positive PCRs considered to be the result of a contaminant (**Figure 2B**). 48/163 (29%, 95% CI 22-36%) positive results led to a change in antimicrobial management. Positive results led to a change in antibiotic management in 5/5 cases (100%) of intervertebral discs and 8/11 (72%, 95% CI 46-98%) of heart valves, while certain specimen types such as liver, brain, and BAL did not have any results lead to a change in patient management. Reasons that antimicrobial management was not changed include the patient already being on appropriate empiric therapy (68/163, 42% [95% CI 34%-50%]), the patient improving without antibiotic intervention (8/163, 5% [95% CI 2-8%]), awaiting mycobacterial susceptibility results from a concurrently submitted culture (2/163, 2% [95% CI 0-4%]), or patient discharge to hospice or death prior to return of the results (4/163, 3% [95% CI 1-5%]).

**Figure 2.**
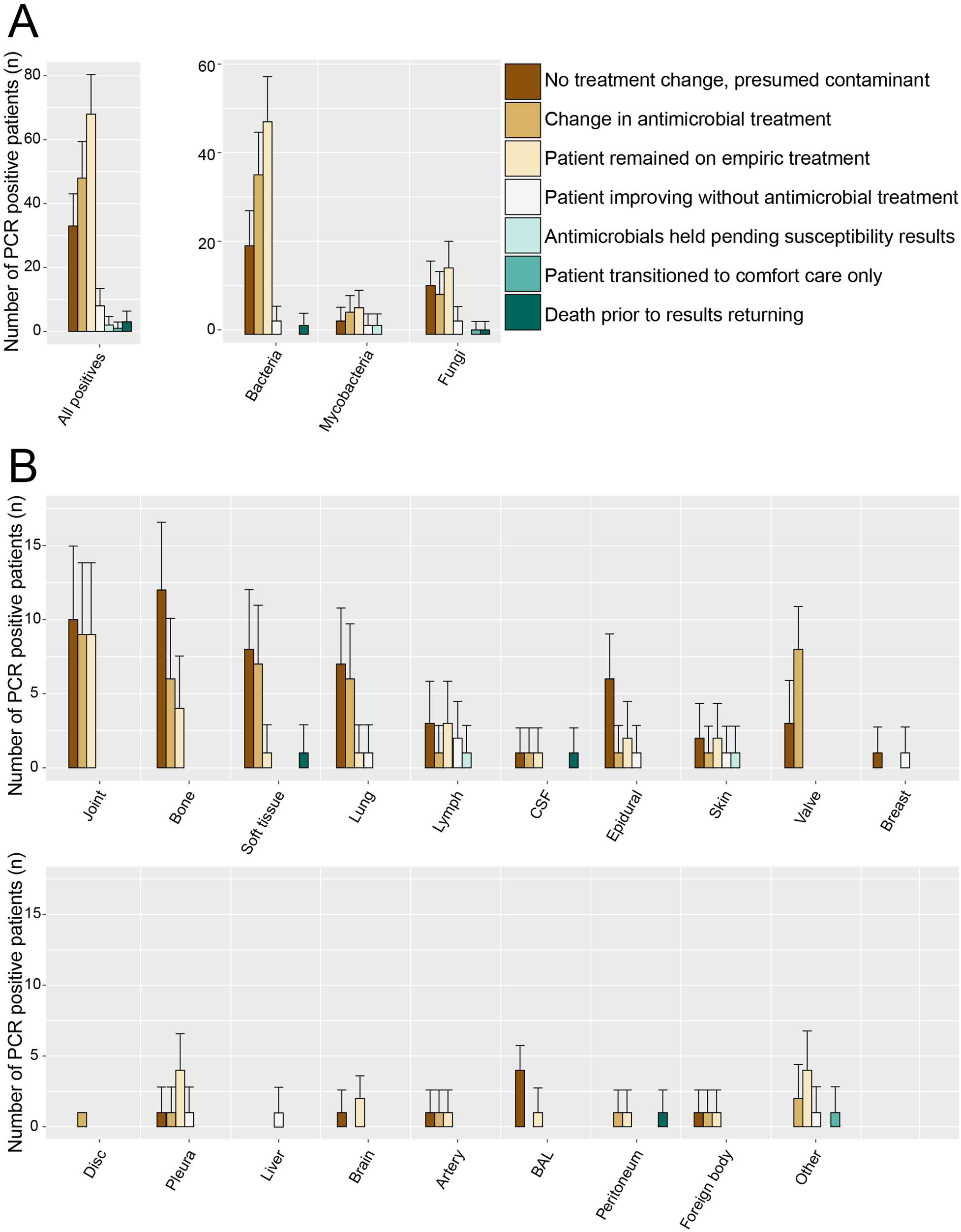
Management decisions made by the clinical team based on positive universal PCR results by pathogen type (A) and by specimen source (B). Error bars represent the 95% population proportion confidence intervals for positive results. Antimicrobial course was not changed in the case of death, transition to comfort care, patient improvement without intervention, satisfactory empiric treatment, or presumed contamination of universal PCR sample.

Finally, we determined whether a positive universal PCR result was associated with particular hospitalization outcomes. Of the 708 total specimens, 593 were from inpatients and 115 were from outpatients, with 136/593 (23%, 95% CI 16-31%) and 27/115 (23%, 95% CI 20-26%) being positive, respectively. From the time of specimen collection, universal PCR results from inpatients took an average of 12 days to return, and 76% (95% CI 72%-79%) of the results returned after the patient was discharged from the hospital. Among inpatients who were tested via universal PCR, 13/593 (2%, 95% CI 1-4%) died, 204/593 (34%, 95% CI 31-38%) required discharge to extended care facilities, including rehabilitation services and hospice care, and 123/593 (21%, 95% CI 17-24%) were readmitted to the hospital within 30 days of discharge. Universal PCR results returning prior to hospital discharge did not significantly affect hospitalization outcomes such as mortality and discharge to rehab services compared to patients whose results did not return until after discharge.

A positive universal PCR for fungi was significantly associated with increased hospital mortality (OR 7.65 [95% CI 1.80 – 32.4], p <0.05) (**Figure 3**). A positive fungal PCR result was not significantly associated with discharge to rehabilitation or hospice care facilities (OR 2.38 [95% CI 1.10-5.18, p=0.052) or readmission within 30 days following hospital discharge (OR 2.09 [95% CI 0.93-4.76], p=0.07). There were no significant differences in hospital mortality, discharge to rehabilitation or hospice facilities, or hospital readmission within 30 days among inpatients with positive bacterial or mycobacterial results. There were no significant differences in hospitalization length of stay associated with positive results for bacteria, mycobacteria, or fungi **(Supplemental Figure 1)**.

**Figure 3.**
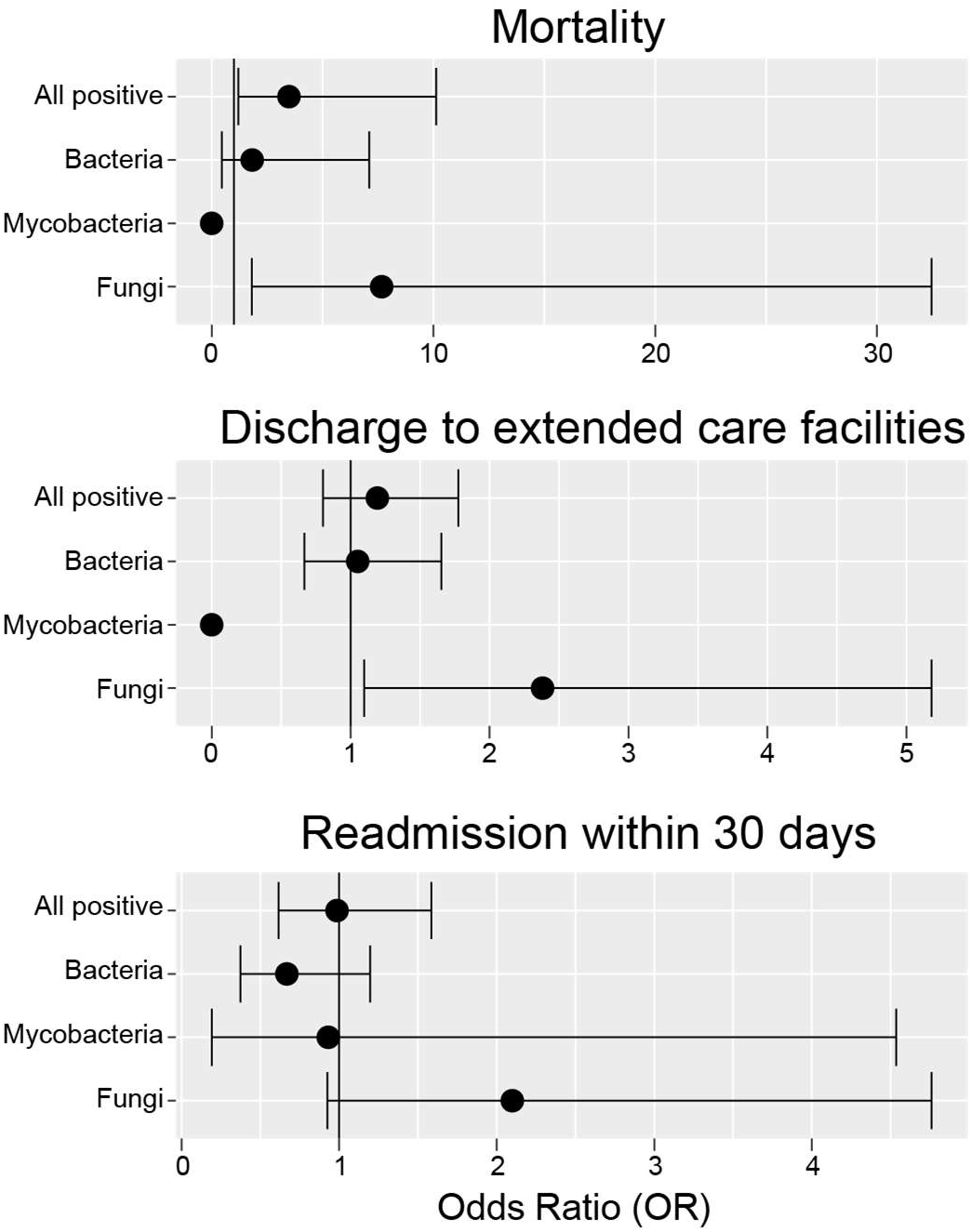
Hospitalization outcomes following positive universal PCR results compared to specimens which were negative via universal PCR. Error bars represent the 95% confidence interval around the odds ratio.

## Discussion

In this study, we examined clinical factors associated with positive universal PCR results, as well as associated hospitalization outcomes. In line with prior literature, we found that positive Gram stain results or histology results, neutrophils on Gram stain, and growth in culture were all significantly associated with positive universal PCR results^25, 31^.

Our clinicians ordered universal PCR on a wide variety of specimen types, most frequently in the case of orthopedic infections. We found that although they were the most frequently ordered, orthopedic specimens such as joint, bone, and intervertebral discs were among the least likely specimens to return positive. While less frequently ordered, heart valves were among the highest positivity rates and most frequently led to changes in antibiotic management. These findings are in line with current literature demonstrating great utility of universal PCR for endocarditis, so much so that some studies have even suggested that universal PCR be incorporated as part of the modified Duke’s criteria for diagnosis of infective endocarditis^22^.

In our cohort, 20% of all positive universal PCR results were considered contaminants by clinical teams. We relied on clinical judgement about whether a positive universal PCR result represented a true infection or contamination. We believe that the fraction of results considered “contaminated” was actually underestimated in our data set due to clinicians potentially over-treating clinically insignificant organisms detected on universal PCR out of an abundance of caution (Note, PCR disagreed with culture-based identification in 32% of cases). With a lack of gold standard in determining whether a resulting organism is truly causative of infection, universal PCR will likely continue to produce organism identifications that will present a clinical dilemma over whether to treat.

Among patients with positive universal PCR results, only 29% of results led to a change in antibiotic management. The impact of positive universal PCR results on antibiotic management varies widely in the literature, with studies showing a change in management in 5% to 76% of cases^16, 24, 25, 32^. The majority of our patients were on empiric treatment prior to universal PCR results, and so clinicians must consider the value that universal PCR brings to potentially changing or narrowing treatment, and how strongly that will impact their patients’ care. A limitation of this study is that we were unable to perform a similar analysis to determine the changes in antibiotic management that may have resulted from a negative universal PCR result. While positive universal PCR results were invariably acknowledged in clinician notes, negative results were less frequently documented and therefore could not be directly correlated with treatment decisions. This may be a fruitful topic for future work.

We found that positive results from universal PCR for fungi were associated with worse hospital outcomes. 4/30 (13%) positive cases in hospitalized inpatients were associated with patient death, and 15/30 (50%) cases required discharge to extended care facilities or hospice. This finding likely reflects that patients with occult fungal infections are likely to be sicker with more severe systemic illness compared to patients without infections as the etiology of their disease.

There were no circumstances in which a positive result of universal PCR led to demonstrable improvements in hospitalization outcomes. Patients with positive universal PCR results for bacteria and mycobacteria did not have any change in hospital mortality, discharge to extended care facilities, readmission within 30 days, or length of stay. However, our universal PCR results took an average of 12 days from specimen collection to result, and 76% of hospitalized patients were discharged before results returned. When considering the utility of this assay, turnaround time is a critical component. With improvements in turnaround time through the development of commercialized assays, it is possible that earlier time to appropriate antimicrobial intervention may be able to improve hospitalization outcomes^9, 13^. However, many of these patients were treated empirically with broad-range antibiotics prior to results and it can be difficult to parse out specific improvements that occurred as a result of potentially narrowing coverage. It can be challenging to quantify other metrics of improper antibiotic use such as medication side effects, excess healthcare cost, and the promotion of resistance based on a retrospective analysis. With that said, universal PCR data may also prove valuable to infection control for epidemiologic studies; for example, in our hospital network, a cluster of four cases of *Acinetobacter junii* detected in orthopedic specimens in 2022 prompted an internal infection control investigation to identify potential sources of OR contamination to reduce false positive rates.

Further studies focused on other outcomes as outlined above are needed to fully assess the utility of universal PCR for the diagnosis of occult infections. With little changes in hospitalization outcomes occurring as a result of universal PCR, clinicians should carefully consider the desired outcomes of a universal PCR weighed against the potential high cost of testing and the risk of over-treating organisms which may not truly be causative of infection. Given the significant variation in ordering and treatment practices among physicians, there will continue to be fluctuations in positivity rates and changes in antimicrobial practice from institution to institution until standardized guidelines are established for the use of this technology, highlighting the need for intensive studies into the utility of universal PCR in clinical practice.

## Data Availability

All data produced in the present study are available upon reasonable request to the authors

## Acknowledgements

We thank the University of Washington Molecular Diagnosis Microbiology Section (Seattle, WA), our reference laboratory to whom we sent all samples to perform universal PCR studies. We also thank our clinical and laboratory staff for specimen submission and processing.

## Author Contributions

J.K. and S.R. conceived the studies and established parameters for data collection. A.M. and R.A. retrieved the data from our institution’s clinical data repository. Data analysis was performed by J.K. and A.K. Manuscript was written by J.K, R.A, and S.R. All authors reviewed the manuscript, data, and approved the final version.

## References

1. Church DL, Cerutti L, Gürtler A, Griener T, Zelazny A, Emler S. Performance and Application of 16S rRNA Gene Cycle Sequencing for Routine Identification of Bacteria in the Clinical Microbiology Laboratory. Clin Microbiol Rev. 2020;33(4):e00053–19. doi:10.1128/CMR.00053-19

2. Sontakke S, Cadenas MB, Maggi RG, Diniz PPVP, Breitschwerdt EB. Use of broad range16S rDNA PCR in clinical microbiology. Journal of Microbiological Methods. 2009;76(3):217–225. doi:10.1016/j.mimet.2008.11.002

3. Morel AS, Dubourg G, Prudent E, et al. Complementarity between targeted real-time specific PCR and conventional broad-range 16S rDNA PCR in the syndrome-driven diagnosis of infectious diseases. Eur J Clin Microbiol Infect Dis. 2015;34(3):561–570. doi:10.1007/s10096-014-2263-z

4. Rampini SK, Bloemberg GV, Keller PM, et al. Broad-Range 16S rRNA Gene Polymerase Chain Reaction for Diagnosis of Culture-Negative Bacterial Infections. Clinical Infectious Diseases. 2011;53(12):1245–1251. doi:10.1093/cid/cir692

5. Schuurman T, de Boer RF, Kooistra-Smid AMD, van Zwet AA. Prospective Study of Use of PCR Amplification and Sequencing of 16S Ribosomal DNA from Cerebrospinal Fluid for Diagnosis of Bacterial Meningitis in a Clinical Setting. J Clin Microbiol. 2004;42(2):734–740. doi:10.1128/JCM.42.2.734-740.2004

6. Tkadlec J, Peckova M, Sramkova L, et al. The use of broad-range bacterial PCR in the diagnosis of infectious diseases: a prospective cohort study. Clinical Microbiology and Infection. 2019;25(6):747–752. doi:10.1016/j.cmi.2018.10.001

7. Lass-Flörl C, Mutschlechner W, Aigner M, et al. Utility of PCR in Diagnosis of Invasive Fungal Infections: Real-Life Data from a Multicenter Study. J Clin Microbiol. 2013;51(3):863–868. doi:10.1128/JCM.02965-12

8. Deutch S, Pedersen LN, Pødenphant L, et al. Broad-range real time PCR and DNA sequencing for the diagnosis of bacterial meningitis. Scandinavian Journal of Infectious Diseases. 2006;38(1):27–35. doi:10.1080/00365540500372861

9. Welinder-Olsson C, Dotevall L, Hogevik H, et al. Comparison of broad-range bacterial PCR and culture of cerebrospinal fluid for diagnosis of community-acquired bacterial meningitis. Clinical Microbiology and Infection. 2007;13(9):879–886. doi:10.1111/j.1469-0691.2007.01756.x

10. Mishra D, Satpathy G, Chawla R, Venkatesh P, Ahmed NH, Panda SK. Utility of broad-range 16S rRNA PCR assay versus conventional methods for laboratory diagnosis of bacterial endophthalmitis in a tertiary care hospital. Br J Ophthalmol. 2019;103(1):152–156. doi:10.1136/bjophthalmol-2018-312877

11. Ogawa M, Sugita S, Watanabe K, Shimizu N, Mochizuki M. Novel diagnosis of fungal endophthalmitis by broad-range real-time PCR detection of fungal 28S ribosomal DNA. Graefes Arch Clin Exp Ophthalmol. 2012;250(12):1877–1883. doi:10.1007/s00417-012-2015-7

12. Fenollar F, Roux V, Stein A, Drancourt M, Raoult D. Analysis of 525 Samples To Determine the Usefulness of PCR Amplification and Sequencing of the 16S rRNA Gene for Diagnosis of Bone and Joint Infections. J Clin Microbiol. 2006;44(3):1018–1028. doi:10.1128/JCM.44.3.1018-1028.2006

13. Kühn C, Disqué C, Mühl H, Orszag P, Stiesch M, Haverich A. Evaluation of Commercial Universal rRNA Gene PCR plus Sequencing Tests for Identification of Bacteria and Fungi Associated with Infectious Endocarditis. J Clin Microbiol. 2011;49(8):2919–2923. doi:10.1128/JCM.00830-11

14. Gille J, Wallstabe S, Schulz AP, Paech A, Gerlach U. Is non-union of tibial shaft fractures due to nonculturable bacterial pathogens? A clinical investigation using PCR and culture techniques. J Orthop Surg Res. 2012;7(1):20. doi:10.1186/1749-799X-7-20

15. Qu X, Zhai Z, Li H, et al. PCR-Based Diagnosis of Prosthetic Joint Infection. J Clin Microbiol. 2013;51(8):2742–2746. doi:10.1128/JCM.00657-13

16. Lampejo T, Ciesielczuk H, Lambourne J. Clinical utility of 16S rRNA PCR in pleural infection. Journal of Medical Microbiology. 2021;70(5). doi:10.1099/jmm.0.001366

17. Insa R, Marín M, Martín A, et al. Systematic Use of Universal 16S rRNA Gene Polymerase Chain Reaction (PCR) and Sequencing for Processing Pleural Effusions Improves Conventional Culture Techniques. Medicine. 2012;91(2):103–110. doi:10.1097/MD.0b013e31824dfdb0

18. Lieberman JA, Bryan A, Mays JA, et al. High Clinical Impact of Broad-Range Fungal PCR in Suspected Fungal Sinusitis. Hanson KE, ed. J Clin Microbiol. 2021;59(11):e00955–21. doi:10.1128/JCM.00955-21

19. Faraji R, Behjati-Ardakani M, Moshtaghioun SM, et al. The diagnosis of microorganism involved in infective endocarditis (IE) by polymerase chain reaction (PCR) and real-time PCR: A systematic review. The Kaohsiung J of Med Scie. 2018;34(2):71–78. doi:10.1016/j.kjms.2017.09.011

20. Harris KA, Yam T, Jalili S, et al. Service evaluation to establish the sensitivity, specificity and additional value of broad-range 16S rDNA PCR for the diagnosis of infective endocarditis from resected endocardial material in patients from eight UK and Ireland hospitals. Eur J Clin Microbiol Infect Dis. 2014;33(11):2061–2066. doi:10.1007/s10096-014-2145-4

21. Armstrong C, Kuhn TC, Dufner M, et al. The diagnostic benefit of 16S rDNA PCR examination of infective endocarditis heart valves: a cohort study of 146 surgical cases confirmed by histopathology. Clin Res Cardiol. 2021;110(3):332–342. doi:10.1007/s00392-020-01678-x

22. Marín M, Muñoz P, Sánchez M, et al. Molecular Diagnosis of Infective Endocarditis by Real-Time Broad-Range Polymerase Chain Reaction (PCR) and Sequencing Directly From Heart Valve Tissue. Medicine. 2007;86(4):195–202. doi:10.1097/MD.0b013e31811f44ec

23. Kerkhoff AD, Rutishauser RL, Miller S, Babik JM. Clinical Utility of Universal Broad-Range Polymerase Chain Reaction Amplicon Sequencing for Pathogen Identification: A Retrospective Cohort Study. Clinical Infectious Diseases. 2020;71(6):1554–1557. doi:10.1093/cid/ciz1245

24. Marbjerg LH, Holzknecht BJ, Dargis R, Dessau RB, Nielsen XC, Christensen JJ. Commercial bacterial and fungal broad-range PCR (Micro-Dx^TM^) used on culture-negative specimens from normally sterile sites: diagnostic value and implications for antimicrobial treatment. Diagnostic Microbiology and Infectious Disease. 2020;97(2):115028. doi:10.1016/j.diagmicrobio.2020.115028

25. Naureckas Li C, Nakamura MM. Utility of Broad-Range PCR Sequencing for Infectious Diseases Clinical Decision Making: a Pediatric Center Experience. McElvania E, ed. J Clin Microbiol. 2022;60(5):e02437–21. doi:10.1128/jcm.02437-21

26. Renvoisé A, Brossier F, Sougakoff W, Jarlier V, Aubry A. Broad-range PCR: Past, present, or future of bacteriology? Médecine et Maladies Infectieuses. 2013;43(8):322–330. doi:10.1016/j.medmal.2013.06.003

27. Aggarwal D, Kanitkar T, Narouz M, Azadian BS, Moore LSP, Mughal N. Clinical utility and cost-effectiveness of bacterial 16S rRNA and targeted PCR based diagnostic testing in a UK microbiology laboratory network. Sci Rep. 2020;10(1):7965. doi:10.1038/s41598-020-64739-1

28. Stempak LM, Vogel SA, Richter SS, Wyllie R, Procop GW. Routine Broad-Range Fungal Polymerase Chain Reaction With DNA Sequencing in Patients With Suspected Mycoses Does Not Add Value and Is Not Cost-Effective. Archives of Pathology & Laboratory Medicine. 2019;143(5):634–638. doi:10.5858/arpa.2017-0299-OA

29. Miller K, Harrington SM, Procop GW. Acid-fast Smear and Histopathology Results Provide Guidance for the Appropriate Use of Broad-Range Polymerase Chain Reaction and Sequencing for Mycobacteria. Archives of Pathology & Laboratory Medicine. 2015;139(8):1020–1023. doi:10.5858/arpa.2013-0705-OA

30. Panousis K, Grigoris P, Butcher I, Rana B, Reilly JH, Hamblen DL. Poor predictive value of broad-range PCR for the detection of arthroplasty infection in 92 cases. Acta Orthopaedica. 2005;76(3):341–346. doi:10.1080/00016470510030805

31. Basein, Tinzar, Gardiner, Bradley, Andujar-Vazquez, Gabriela, et al. Clinical Utility of Universal PCR and its Real-world Impact on Patient Management. In: IDSA Stewardship: Impact of Diagnostics. Vol 4. ; 2017:S627.

32. Racsa LD, DeLeon-Carnes M, Hiskey M, Guarner J. Identification of bacterial pathogens from formalin-fixed, paraffin-embedded tissues by using 16S sequencing: retrospective correlation of results to clinicians’ responses. Human Pathology. 2017;59:132–138. doi:10.1016/j.humpath.2016.09.015

